# Population Impact of a hybrid strategy combining maternal RSV vaccination and nirsevimab immunization on lower respiratory tract infections in infants under 1 year in Bogotá: a counterfactual analysis

**DOI:** 10.64898/2026.07.05.26357339

**Authors:** Julián A. Fernández-Niño, A. Alejandra Marín, Luz. A Gutiérrez, María F. Tovar, Diana M. Ayala, Mary L. Gomez, María B. Jaimes, Marcela Martínez, Patricia E. Molano, Diana S. Ríos, Diana M. Walteros, Gerson O. Bermont

## Abstract

**Objective:** To assess the impact of a hybrid RSV immunization strategy on hospitalizations, pediatric intensive care unit (ICU) admissions, outpatient visits, and mortality due to LRTI among infants under one year of age in Bogotá.

**Methods:** We conducted an ecological interrupted time-series study using weekly surveillance data from Bogotá from EW 1 of 2023 to EW 24 of 2026 (181 weeks). Outcomes included weekly rates of all-cause viral LRTI-related general hospitalizations, pediatric ICU admissions, outpatient visits, and deaths among infants younger than one year. Segmented negative binomial regression models adjusted for secular trends, seasonality using Fourier terms, and autocorrelation were used to estimate changes associated with maternal RSVpreF vaccination and nirsevimab implementation. Counterfactual analyses were performed to estimate cases averted and relative risk reductions.

**Results:** Compared with the same period in 2025, the 2026 LRTI hospitalization rate decreased significantly (rate ratio [RR] 0.66; 95% CI 0.64–0.68), as did pediatric ICU admissions (RR 0.78; 95% CI 0.73–0.85) and outpatient visits (RR 0.78; 95% CI 0.77–0.79). Interrupted time-series analyses identified a significant weekly decline in hospitalization trends following maternal RSVpreF introduction (3.9% per week; p=0.023) and a smaller but significant decline in ICU admissions (−2.8% per week; p=0.039). The cumulative relative reduction in hospitalizations was estimated at 47.1% (95% CI 13.9–70.4), corresponding to 7,605 hospitalizations averted over the post-intervention period (EW 47/2025–EW 24/2026). No statistically significant changes were observed for outpatient visits or mortality.

**Conclusions:** Implementation of a hybrid RSV prevention strategy was associated with a substantial reduction in severe LRTI among infants during the first respiratory season following introduction in Bogotá. These findings provide the first real-world population-level evidence from Latin America supporting hybrid RSV immunization as a feasible and potentially cost-effective strategy for reducing severe infant respiratory disease in middle-income settings.

## Introduction

Respiratory syncytial virus (RSV) is the leading cause of acute lower respiratory tract infection (LRTI) in infants, resulting in substantial morbidity and mortality, particularly among children under five years of age [1]. Severe disease occurs most frequently in infants younger than six months and in those with underlying medical conditions or multiple comorbidities [2]. Globally, RSV is estimated to cause approximately 6.6 million episodes of LRTI each year, leading to about 1.4 million hospitalizations and nearly 45,700 deaths annually among children under six months of age [3], with 97% of these deaths occurring in low- and middle-income countries [3].

In Colombia, RSV remains a major preventable cause of infant morbidity and mortality and an important public health burden [4]. RSV circulates continuously and is among the most prevalent respiratory viruses, accounting for around 32.4% of overall viral detections among children younger than 5 years [5]. In particular, Bogotá has a sustained year-round RSV circulation presenting two seasonal peaks of intense respiratory virus circulation (April–June and September–November) [6]. This epidemiological pattern imposes a substantial burden on the health-care system, particularly on pediatric services [7], as densely populated urban settings with crowding, high population mobility, and shared indoor environments facilitate rapid viral transmission [7]. In 2025, Bogotá reported a cumulative total of 20,470 general hospitalizations and 3,548 admissions to pediatric intensive care units (ICUs) among children younger than 1 year due to LRTI [8]. Of these, 445 laboratory-confirmed RSV cases which represents 52.4 % of all viral detections and were identified through sentinel surveillance, which provides a reliable representation of population-level RSV circulation patterns [9].

Recent advances in immunoprophylaxis have reshaped the landscape of RSV prevention. Long-acting monoclonal antibodies (mAb) such as nirsevimab (Beyfortus®, Sanofi) provide season long protection with a single dose [10,11], representing a major improvement over earlier multidose and high-cost interventions such as palivizumab [12,13]. In parallel, maternal RSV vaccination (Abrysvo®; Pfizer) has emerged as an effective strategy to confer early protection through transplacental antibody transfer [14,15]. Two studies conducted in South America during the 2024 RSV season have demonstrated a substantial reduction in RSV-associated severe outcomes in real-world settings. In Chile, the introduction of a universal infant immunization strategy with the long-acting mAb nirsevimab during the 2024 RSV season showed a 76% of reduction in RSV- LRTI hospitalizations and an approximately 85% of RSV-related intensive care unit admissions, with an estimated 77% relative reduction in RSV-related hospitalizations at the population level compared with a counterfactual no-intervention scenario [16]. Similarly, in Argentina, the nationwide maternal RSV immunization programme showed a reduction in RSV-associated LRTI hospitalizations of approximately 79% in infants up to 3 months of age and 71% up to 6 months [17]. In addition, few countries worldwide have implemented RSV hybrid preventative strategies . Recently, Australia implemented a hybrid RSV prevention strategy, prioritizing maternal vaccination, with nirsevimab reserved for infants born to unvaccinated mothers and selected high-risk groups. The program’s impact showed that the hospitalizations declined most markedly among infants aged 0 to <3 months (43.8%), followed by reductions of 20.1% in those aged 3 to <6 months and 8.5% in infants aged 6 to 12 months[18].

Based on the accumulated real-world evidence related to the potential of both maternal RSV vaccination and nirsevimab immunization to substantially reduce severe RSV disease and alleviate hospital burden during early infancy, Bogotá decided to implement a hybrid strategy starting in November 2025 which combines maternal RSV vaccination (28 to 36 weeks of pregnancy) as the primary approach, and nirsevimab immunization for premature infants and newborns from unvaccinated mothers and high-risk conditions starting in February 2026. This approach was based on cost-effectiveness and multi-criteria analyses carried out by a technical advisory group of the University of Antioquia and District Health Secretariat of Bogotá, as well as providers and patient organizations, who identified nirsevimab and Abrysvo as more cost-effective alternatives compared to palivizumab and a no-intervention scenario. [19–22]

The hybrid strategy represents a feasible and cost-efficient approach for low and middle-income settings, enabling expanded RSV immunization coverage through direct or indirect protection of all newborns, in contrast to universal nirsevimab immunization, which remains economically viable only in high-income countries [23]. Here, we report novel real-world evidence from Latin America of a hybrid immunization strategy that combines maternal RSV vaccination and neonatal immunization, assessing its impact on LRTI outcomes among infants under 1 year of age in Bogotá, providing evidence to other middle-income settings considering combined prevention approaches.

## Methods

### Study design

We conducted an ecological, analytical interrupted time-series (ITS) study to assess the population-level impact of maternal and newborn immunization strategies against RSV on LRTI outcomes among infants younger than 1 year residing in Bogotá, Colombia. The study period spanned from epidemiological week 1 to 24 of 2023 to 1- 24 of 2026, enabling comparison of bimodal epidemiological patterns across pre- and post-intervention periods. This design allowed for the assessment of pre-intervention trends and the estimation of post-intervention changes relative to a counterfactual no-intervention scenario.

### Data sources and population

We included infants aged under 1 year to assess LRTI morbidity, using hospitalization rates weekly aggregated in both general and pediatric ICUs as indicators of disease burden. LRTI was defined as cough or difficulty breathing, onset of symptoms within the preceding 10 days, and at least one of the following: fast breathing (respiratory rate ≥60 breaths per min for infants aged <2 months, respiratory rate ≥50 breaths per min for infants aged 2–6 months); oxygen saturation (SpO2) less than 95%; or chest wall indrawing.

Data were obtained from Colombia’s National Public Health Surveillance System (SIVIGILA), specifically from event 995, which consolidates notifications of ICD-10 diagnosis codes (J00–J22), allowing for the assessment of disease burden and all causes of LRTI. In addition, we used reports from event 591 which captures integrated mortality surveillance among children under five years of age due to LRTI, acute diarrheal disease (ADD), and acute malnutrition (AMN).

### Outcomes

To estimate the population-level impact of the interventions, we defined outcomes using a broad category of LRTI attributable to all-cause viral pathogens, rather than restricting analyses to RSV-specific diagnoses. The primary outcomes were population-based weekly rates of LRTI-related healthcare utilization and mortality among infants aged <1 year, including general hospitalizations, pediatric intensive care unit (ICU) admissions, outpatient visits and deaths attributable to LRTI.

### Statistical analysis

An interrupted time series (ITS) regression with segmented regression was used to estimate the population-level impact of the hybrid immunization strategy on each outcome, following the framework described by Lopez Bernal et al [24] and previously applied to RSV immunization programs by Villa et al [25]. The unit of analysis was the epidemiological week (EW).

Let *Y_t_* be the observed weekly count of an outcome in week *t* (t = 0, …, 180; the 181 consecutive weeks from EW 1 of 2023 to EW 24 of 2026). Counts were modeled with a negative binomial distribution to accommodate overdispersion; the dispersion parameter was estimated using the Cameron–Trivedi method of moments, with a Poisson fit as an auxiliary step. The expected weekly count was modeled on the logarithmic scale, as summarized in equation 1 (see the supplementary table 1).

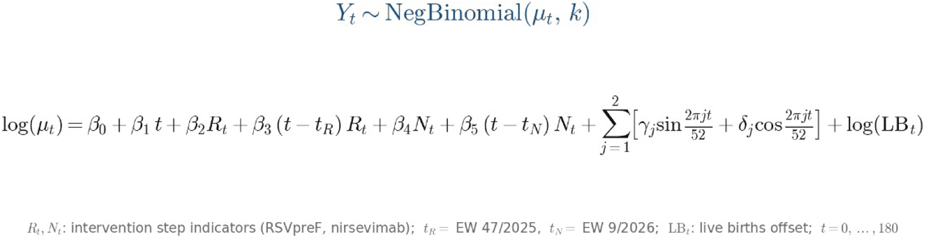

This model separates the effect of each intervention into two complementary components. The level change (β_2_ for RSVpreF, β_4_ for nirsevimab) is the immediate, step-like shift in the rate at the moment the intervention takes effect, and is consistent with a rapid mechanism of action. The trend change (β_3_ for RSVpreF, β_5_ for nirsevimab) is the change in the weekly slope after the intervention, and captures effects that accumulate gradually as successive birth cohorts become protected. Exponentiating a coefficient yields a multiplicative rate ratio; for example, exp(β_3_) is the weekly relative change in the rate attributable to the trend change, and (exp(β_3_) − 1) × 100 expresses it as a percentage change per week.

### Intervention cut-points

The RSVpreF cut-point was set at EW 47/2025, incorporating a lag of approximately 5 weeks between maternal vaccination (administered between weeks 28 and 36 of gestation) and the transfer of transplacental protection to the newborn. The nirsevimab cut-point was set at EW 9/2026, without a lag, since the monoclonal antibody confers immediate passive protection. Because the two cut-points are close in time, the nirsevimab terms were also omitted in a reduced specification (Model A) to bound the contribution of the maternal vaccine in isolation.

### Modeling of year-over-year seasonality

Bogotá exhibits a bimodal pattern of RSV circulation, with two seasonal peaks per year (approximately April–June and September–November) superimposed on sustained year-round transmission. To capture this recurring year-over-year seasonality without consuming one degree of freedom for each calendar week, it was represented by pairs of Fourier harmonic terms (sine and cosine functions) of the annual cycle [24,26]. The first harmonic (52-week period) models the dominant annual wave, and the second harmonic (26-week period) accommodates the secondary semiannual peak that gives the series its bimodal shape. Two harmonic pairs (periods of 52 and 26 weeks) were specified to represent this bimodal annual cycle, and the adequacy of this choice was examined in sensitivity analyses by varying the number of harmonics. Because they are smooth periodic functions of time, these terms share the same seasonal shape across years, which allows the segmented terms to isolate the intervention effect from the expected seasonal fluctuation.

### Denominator, inference, and counterfactual estimation

To express results as rates rather than counts and to account for the secular decline in births in Bogotá over the study period, the weekly number of live births from the RUAF registry was included as an offset, log(NV*_t_*). To account for residual serial autocorrelation, Newey–West heteroskedasticity- and autocorrelation-consistent (HAC) robust standard errors were used, with a lag order of 5 for the 179-week series, as in the controlled time series analysis of Otieno et al [26]. The fitted pre-intervention model was projected forward to generate the counterfactual scenario (the expected number of events in the absence of intervention). Population-level impact was quantified as the cumulative number of cases averted (the difference between the counterfactual and the observed series), the relative risk reduction (RRR), each with 95% confidence intervals obtained from 1,000 parametric bootstrap replications.

### Sensitivity analysis

Robustness was examined by varying the number of Fourier harmonics, shifting each cut-point by ±2 weeks, fitting the reduced model without the nirsevimab terms (Model A), and excluding 2023 from the pre-intervention period.

All the analyses were performed in Python (pandas, numpy, statsmodels, and patsy) and R consistent with previous studies of RSV immunization impact [25].

## Results

### Descriptive analysis of the reduction in lower respiratory tract infection burden in infants under 1 year of age

Between epidemiological week (EW) 1 and EW 24 of 2026, a significant decrease was observed in Bogotá in the number and rates of general hospitalizations, ICU admissions, and outpatient visits in 2026 relative to the previous years analyzed since 2023 (Table 1). In particular, the general hospitalization rate was significantly lower compared with the same period in 2025, with a rate ratio (RR) of 0.66 (95% CI: 0.64–0.68; p<0.001). ICU admissions and outpatient visits, in turn, showed a smaller reduction, with an RR of 0.78 (95% CI: 0.73–0.85; p<0.001) and an RR of 0.78 (95% CI: 0.77–0.79; p<0.001), respectively. In addition, during the 2026 study period six deaths from lower respiratory tract infections were reported, of which only three were associated with and confirmed as RSV. The three cases occurred in infants who did not have the opportunity to benefit from full implementation of the hybrid strategy. Two were children born before the start of maternal RSV vaccination and therefore had no possibility of receiving transplacental protection. The third case occurred in an infant born preterm at 32 weeks of gestation, the son of a vaccinated mother, who received a dose of palivizumab in accordance with current recommendations; however, he was born before nirsevimab became available in the city and therefore could not access the neonatal component of the hybrid strategy. Consequently, these events should be interpreted in the context of the staggered implementation period of the strategy and not as evidence of failure of the preventive interventions (Table 1).

**Table 1.**
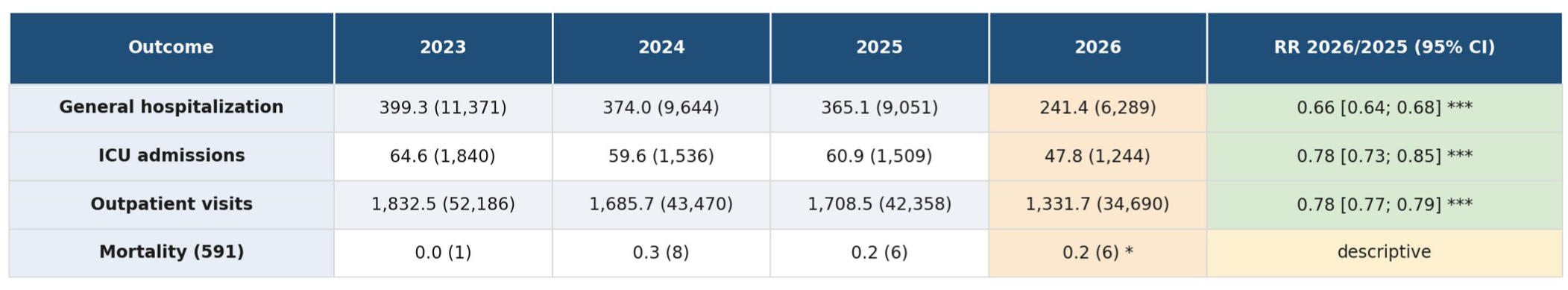
Burden of lower respiratory tract infection in infants under 1 year of age: rates per 1,000 live births, counts, and rate ratios (RR) before and after the implementation of maternal RSVpreF vaccination and nirsevimab immunization. Bogota, D.C., epidemiological weeks (EW) 1-24, 2023-2026. Each cell: rate per 1,000 live births (count in parentheses). Live births (EW 1-24): 2023=28,478 • 2024=25,787 • 2025=24,792 • 2026=26,049. *\*\*\* p<0.001. Crude RR by rate ratio (95% Cl, log-Wald). * Mortality (event 591) in 2026 confirmed only through EW 21 (reporting lag); shown descriptively.*

### Population-level impact of the hybrid immunization strategy on lower respiratory tract infections in infants under 1 year of age

Figure 1 shows the observed time series and the estimated counterfactual scenarios for general hospitalization, pediatric ICU admission, and outpatient visits for viral lower respiratory tract infection in infants under one year of age in Bogotá. During the period prior to implementation of the hybrid immunization strategy, the observed trends followed the expected seasonal pattern. However, after the introduction of maternal RSVpreF vaccination, the observed trajectories began to diverge progressively from those predicted by the counterfactual models, particularly for the general hospitalization outcome.

**Figure 1.**
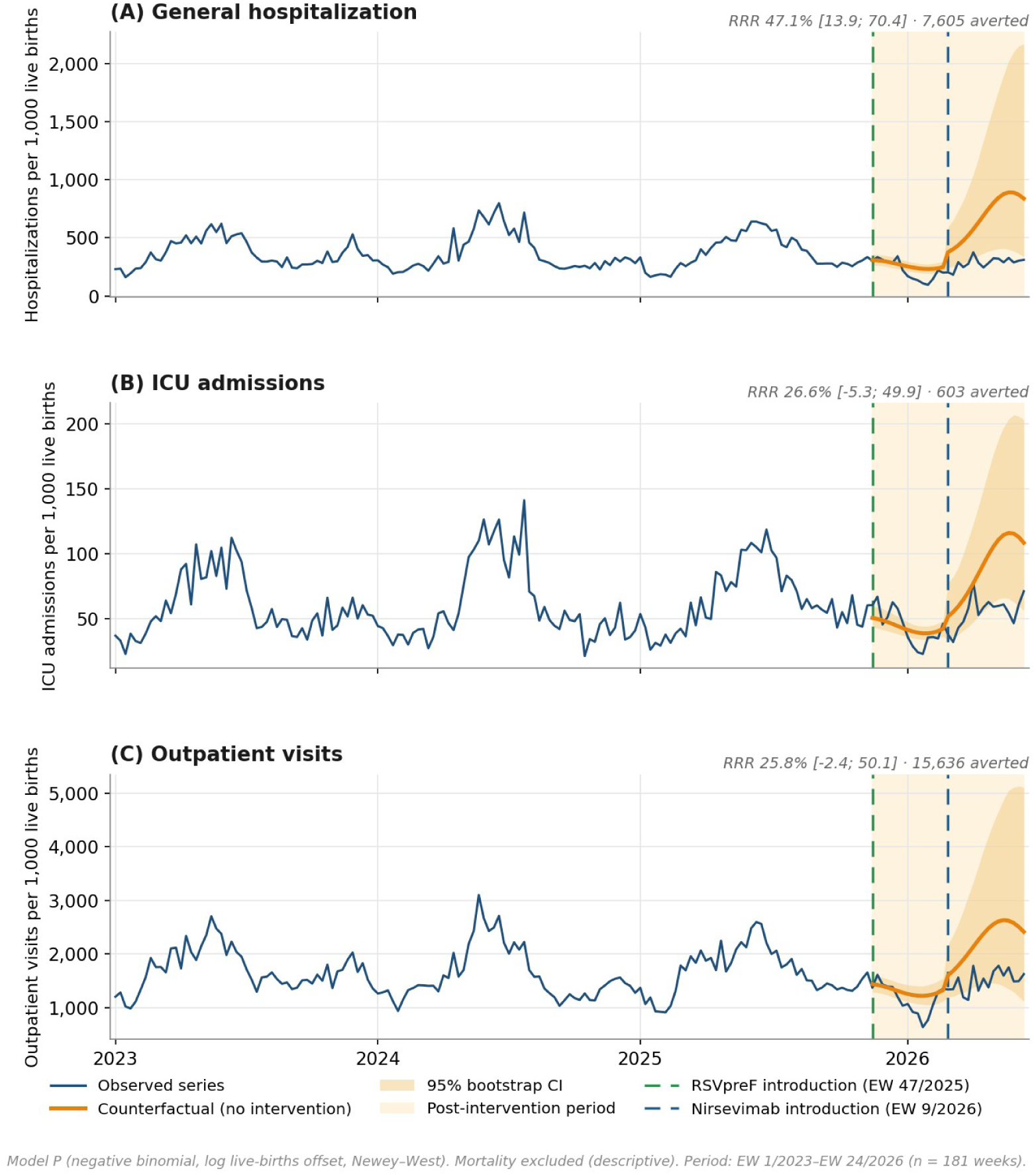
Observed and counterfactual interrupted time series of lower respiratory tract infection (LRTI) outcomes among infants aged <1 year in Bogotá D.C., 2023–2026. (A) General hospitalization. (B) Pediatric intensive care unit (ICU) admission. (C) Outpatient visits. Solid lines represent observed weekly rates, whereas dashed lines represent the counterfactual rates estimated using interrupted time-series models following the introduction of maternal RSVpreF vaccination and nirsevimab immunization.

The ITS model results confirmed that the main effect associated with the intervention corresponded to a sustained reduction in hospitalizations. Although no significant immediate change in the level of the series was observed after the introduction of RSVpreF (+6.5%; 95% CI: −16.8 to 36.3; p=0.615), we identified a significant decline in the trend equivalent to 3.9% per week (95% CI: −7.1 to −0.6; p=0.023) (Table 2). In Figure 1A, this change is reflected in a progressive divergence between the observed series and the counterfactual scenario, which widens as the expected respiratory peak season progresses.

**Table 2.**
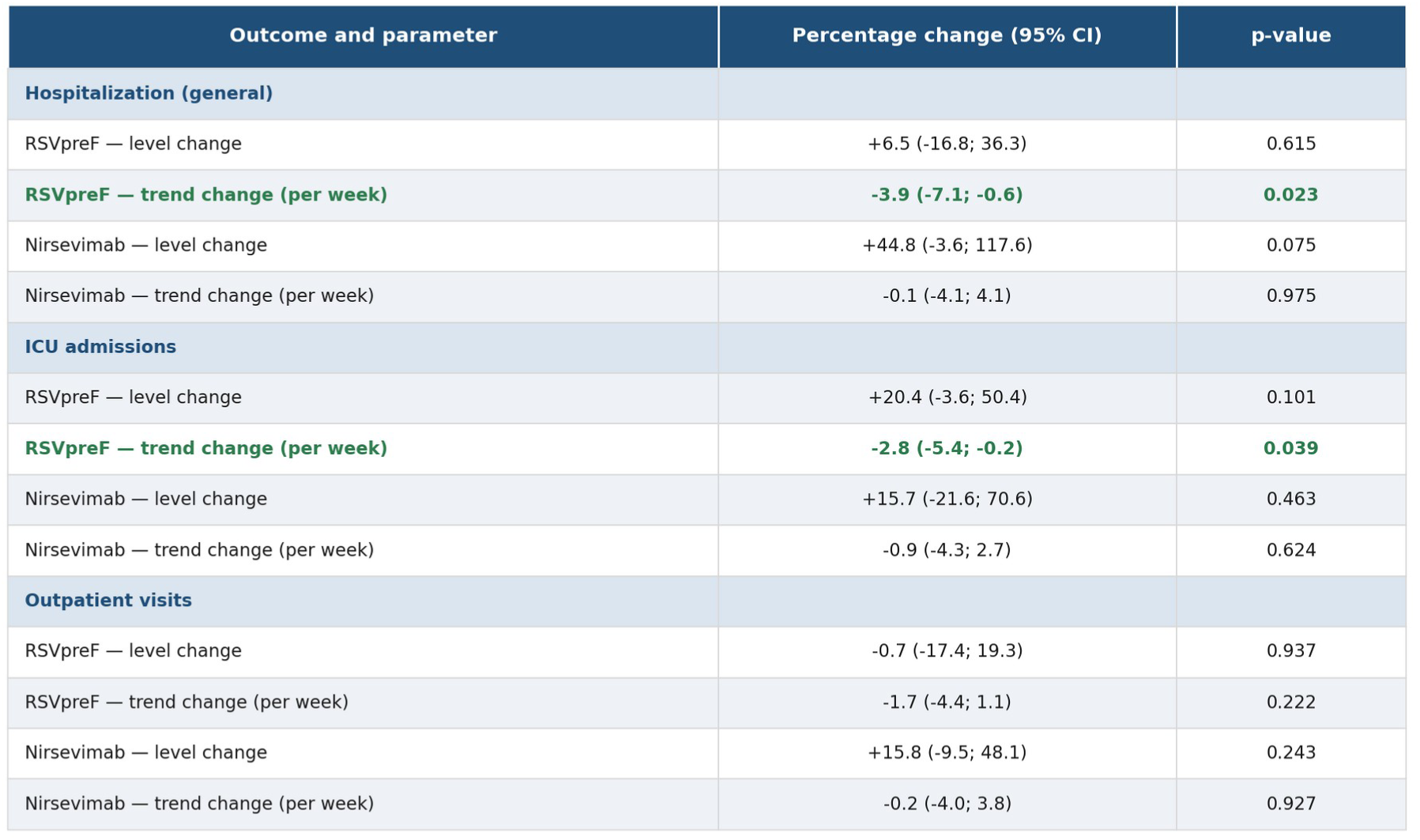
Effect of maternal RSVpreF vaccination and nirsevimab immunization on lower respiratory tract infection outcomes in infants under 1 year of age: interrupted time series analysis. Bogota, D.C., epidemiological weeks (EW) 1/2023-24/2026 (n = 181 weeks). Negative binomial with logflive births) offset, Fourier 52+26 and Newey-West (5 lags). Cut-points: RSVpreF EW 47/2025, nirsevimab EW 9/2026. *Bold = p<0.05. Level = immediate step; trend = change in weekly slope.*

| Outcome and parameter | Percentage change (95% CI) | p-value |
| --- | --- | --- |
| **Hospitalization (general)** |  |  |
| RSVpreF — level change | +6.5 (-16.8; 36.3) | 0.615 |
| **RSVpreF — trend change (per week)** | **-3.9 (-7.1; -0.6)** | **0.023** |
| Nirsevimab — level change | +44.8 (-3.6; 117.6) | 0.075 |
| Nirsevimab — trend change (per week) | -0.1 (-4.1; 4.1) | 0.975 |
| **ICU admissions** |  |  |
| RSVpreF — level change | +20.4 (-3.6; 50.4) | 0.101 |
| **RSVpreF — trend change (per week)** | **-2.8 (-5.4; -0.2)** | **0.039** |
| Nirsevimab — level change | +15.7 (-21.6; 70.6) | 0.463 |
| Nirsevimab — trend change (per week) | -0.9 (-4.3; 2.7) | 0.624 |
| **Outpatient visits** |  |  |
| RSVpreF — level change | -0.7 (-17.4; 19.3) | 0.937 |
| RSVpreF — trend change (per week) | -1.7 (-4.4; 1.1) | 0.222 |
| Nirsevimab — level change | +15.8 (-9.5; 48.1) | 0.243 |
| Nirsevimab — trend change (per week) | -0.2 (-4.0; 3.8) | 0.927 |

By contrast, the introduction of nirsevimab showed no statistically significant changes either in the level (+44.8%; 95% CI: 3.6 to 117.6; p=0.075) or in the trend of hospitalizations (−0.1% per week; 95% CI: −4.1 to 4.1; p=0.975) (Table 2). The absence of a detectable independent effect was probably influenced by the short follow-up period available after its introduction and by the temporal overlap with the effects of RSVpreF.

For pediatric ICU admissions, an immediate increase in the level of the series was observed after the introduction of RSVpreF (+20.4%; 95% CI: −3.6 to 50.4; p=0.101), accompanied by a significant decline in the trend of 2.8% per week (95% CI: −5.4 to −0.2; p=0.039). Visually, the observed series shows a gradual decrease relative to the counterfactual from the weeks following the intervention onward, although with greater variability than that observed for general hospitalization (Figure 1B).

For outpatient visits, the observed and counterfactual curves showed greater overlap throughout the post-intervention period (Figure 1C). Consistently, no significant changes were identified in association with RSVpreF, either in the level (−0.7%; 95% CI: −17.4 to 19.3; p=0.937) or in the trend (−1.7% per week; 95% CI: −4.4 to 1.1; p=0.222), nor with the introduction of nirsevimab (Table 2).

The findings observed in the time series indicated a cumulative relative reduction in general hospitalizations of 47.1% (95% CI: 13.9–70.4), which remained robust in the sensitivity analyses (range 32–47%). This corresponds to 7,605 general hospitalizations averted (95% CI: 1,376–20,282). In addition, the cumulative relative reduction for pediatric ICU admissions was 26.6% (95% CI: −5.3–49.9), with 603 averted cases (95% CI: −84 to 1,658). Finally, for outpatient visits a cumulative relative reduction of 25.8% (95% CI: −2.4–50.1) was observed, with 15,636 averted cases (95% CI: −1,036 to 45,026) over the post-intervention period (EW 47/2025–EW 24/2026) (Table 3).

**Table 3.**
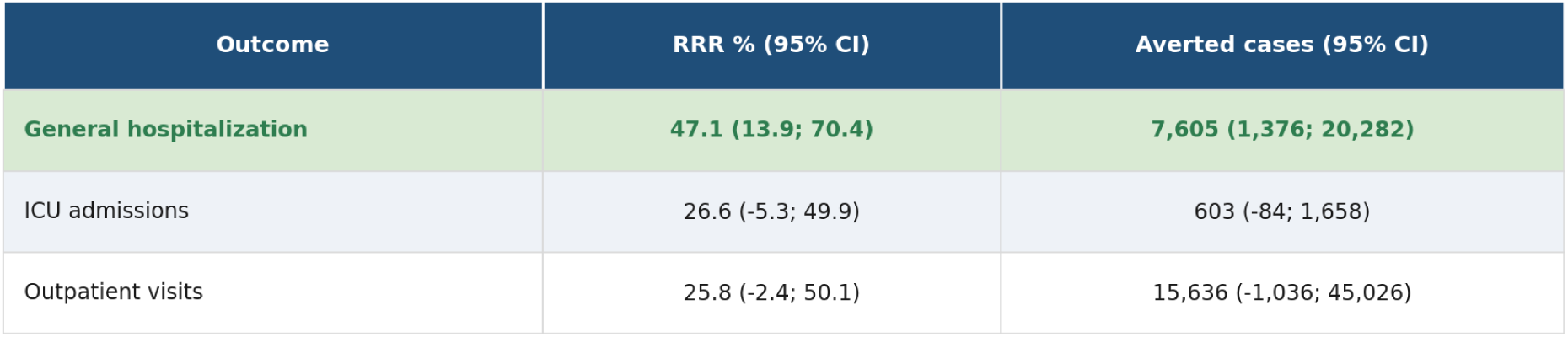
Cumulative relative risk reduction and averted cases of lower respiratory tract infections among infants aged <1 year after the implementation of maternal RSVpreF vaccination and nirsevimab immunization. Bogota, D.C., epidemiological weeks (EW) 47/2025-24/2026. imulative over the post-intervention period, from RSVpreF introduction (EW 47/2025) through EW 24/2026, versus a no-intervention counterfactual; 95% Cis by parametric bootstrap (1,000 replicate: Bold indicates a statistically significant reduction (95% Cl excludes 0). Mortality (event 591) is not shown (confirmed only through EW 21; treated descriptively).

| Outcome | RRR % (95% CI) | Averted cases (95% CI) |
| --- | --- | --- |
| General hospitalization | **47.1 (13.9; 70.4)** | **7,605 (1,376; 20,282)** |
| ICU admissions | 26.6 (-5.3; 49.9) | 603 (-84; 1,658) |
| Outpatient visits | 25.8 (-2.4; 50.1) | 15,636 (-1,036; 45,026) |

## Discussion

This study presents the first real-world evidence of the population-level impact of a hybrid RSV prevention strategy in Bogotá, based on the complementary implementation of maternal RSVpreF vaccination and neonatal nirsevimab immunization. The results show a significant reduction in the historical trend of hospitalizations for acute lower respiratory tract infection in infants under one year of age, as well as in pediatric intensive care unit admissions across the city, which translates into a substantial decrease in disease burden during the first respiratory peak following the introduction of the strategy. Although a reduction in outpatient visits was also observed, it was not statistically significant. In absolute terms, the estimated impact corresponds to a substantial reduction in the hospitalization rate and in severe cases averted from lower respiratory tract infections in infants under one year of age, with potential expected implications for the reduction of medium- and long-term complications and sequelae in early childhood.

This population-level effect can be explained by the complementary nature of the two immunological interventions, each of which has previously demonstrated, independently, high efficacy in clinical trials [10,11] [14,15], as well as effectiveness in real-world conditions [16] [17] [18]. The joint implementation in Bogotá made it possible to extend protection to virtually all newborns, whether through passive immunity conferred by maternal vaccination or through the direct administration of monoclonal antibodies during the neonatal period. This impact was enhanced by the high coverage achieved in Bogotá from the outset of the strategy, where the proportion of newborns of vaccinated mothers increased progressively from the start of the strategy and exceeded 80% during the 2026 respiratory peak. Likewise, the availability of nirsevimab for premature infants and high-risk infants or those of unvaccinated mothers helped to reduce protection gaps in the most vulnerable groups. An additional determinant that contributed to the observed impact was the timeliness of the strategy’s implementation. Maternal vaccination began in late November 2025, approximately three months before the period of highest RSV circulation in Bogotá, which allowed most infants to be born with passive protection during the first respiratory peak, which historically occurs between March and June. Consequently, a high proportion of newborns entered the season of highest viral transmission with maternal antibodies, while those not protected through this route were eligible to receive nirsevimab, thereby maximizing the population coverage achieved by the hybrid strategy [27].

In the absence of new preventive or therapeutic interventions relevant to RSV, or to severe respiratory infections in general, during the study period in Bogotá, it is reasonable to attribute a substantial proportion of the observed effect to the implemented hybrid strategy, especially in the absence of competing causes. This assumption is consistent with the methodological principles of interrupted time-series studies, in which the absence of concurrent interventions strengthens the causal inference attributable to the evaluated intervention [24]. Beyond being plausible, these findings are consistent with international evidence that has demonstrated substantial reductions in pediatric hospitalizations following the introduction of RSV immunization strategies in different global contexts. Chile, for example, through its universal nirsevimab strategy, found a reduction of 76% in RSV-associated lower respiratory tract infection hospitalizations and an approximately 85% reduction in RSV-associated pediatric intensive care admissions [16]. Similarly, in Argentina, the maternal RSV vaccination program showed a 79% reduction in RSV-associated lower respiratory tract infection hospitalizations in infants up to 3 months of age and a 71% reduction in infants up to 6 months of age [17]. By contrast, Australia, through its implementation of a hybrid RSV prevention program, showed a significantly more pronounced impact on the hospitalization rate in infants under 3 months of age, reporting a 43.8% reduction [18]. Although the magnitude of the impact may vary depending on seasonality and the epidemiological dynamics of viral circulation, unlike regions with marked winter seasonality, Bogotá exhibits sustained RSV circulation throughout the year with two epidemic peaks [6], which makes it especially relevant to evaluate interventions in settings of continuous transmission. In this context, the present study provides original evidence on the population-level impact of a hybrid strategy and as a viable and cost-effective alternative for low- and middle-income countries. The successful implementation of this strategy can also be attributed to the strong commitment and dedication of health-care personnel in Bogotá, including vaccinators, obstetricians, and pediatricians, whose role was essential to ensure timely implementation, achieve high coverage, generate demand for the vaccine, educate mothers and caregivers, and ensure adequate operational execution of the intervention.

From a methodological standpoint, this study is strengthened by the use of a population-level interrupted time-series design, with modeling based on negative binomial regression adjusted for secular trend, seasonality through Fourier harmonics, and control of autocorrelation through robust standard errors. The estimation of cumulative impact against a no-intervention counterfactual scenario, together with multiple sensitivity analyses, reinforces the robustness of the findings. [24]

However, important limitations must also be considered. First, the post-implementation evaluation window is currently short (through epidemiological week 24 of 2026), which reduces the statistical power for analyses of the more slowly evolving outcomes; consequently, the cumulative estimates for ICU admissions —whose declining trend did reach statistical significance—, outpatient visits, and mortality were not significant, a pattern compatible with the lower power of a short window and one that should not necessarily be interpreted as absence of effect. These results should be considered preliminary until a complete year-round season is available. Nonetheless, the window was strategic from an analytical standpoint, as it corresponds to the first respiratory peak following implementation —historically the period of most intense RSV circulation in Bogotá— and therefore the one with the greatest signal for detecting population-level impact. Second, the ecological design precludes individual-level linkage: it estimates the population-level impact of the strategy rather than the specific effectiveness of each product, so its results are not directly transferable to the individual level (ecological fallacy). Third, and relatedly, the design does not allow the contribution of maternal RSVpreF vaccination and nirsevimab to be separated, as their introductions were close in time and therefore collinear; nor does it allow the isolation of any residual contribution of palivizumab. Nonetheless, it should be noted that palivizumab is restricted to infants born before 33 weeks of gestation —a small, high-risk subgroup— and was not recorded in the available databases, so its contribution to the population-level effect would be, at best, minimal. Finally, an important limitation of this study is the impossibility of specifically quantifying the population-level impact of the hybrid strategy on RSV-attributable hospitalizations, because city-level information linking the evaluated outcomes to laboratory-confirmed diagnoses was not available. Consequently, future studies incorporating systematic etiological confirmation at the population level will be needed to directly estimate the specific impact of the hybrid strategy on RSV-associated disease burden.

Despite these limitations, this study represents a relevant contribution, as it is, to the best of our knowledge from the available literature, one of the first evaluations of the population-level impact of a hybrid RSV prevention strategy in Latin America. Its results have direct implications for public health, in a context in which multiple countries are currently considering the adoption of RSV immunization strategies. The hybrid strategy emerges as a potentially viable alternative, especially for low- and middle-income countries, by making it possible to extend protection of newborns through the combination of maternal vaccination and targeted neonatal immunization. This approach could optimize the use of available resources by offering broad population coverage at a potentially lower cost compared with strategies based on universal nirsevimab immunization [28].

In conclusion, the implementation of a hybrid strategy based on maternal RSVpreF vaccination and nirsevimab immunization was associated with a significant reduction in the population-level burden of acute lower respiratory tract infection in infants under one year of age during the first respiratory peak following its introduction in Bogotá. These findings support its potential as a public health strategy in middle-income settings and underscore the need to continue the analysis over longer periods, as well as to complement these analyses with individual-level effectiveness studies, future studies with etiological confirmation capacity, and evaluations of other important cost-effectiveness outcomes, to more adequately guide decision-making regarding the incorporation of hybrid RSV prevention strategies at the national and international levels.

## Data Availability

All data produced in the present work are contained in the manuscript

---

**Supplementary table 1.**
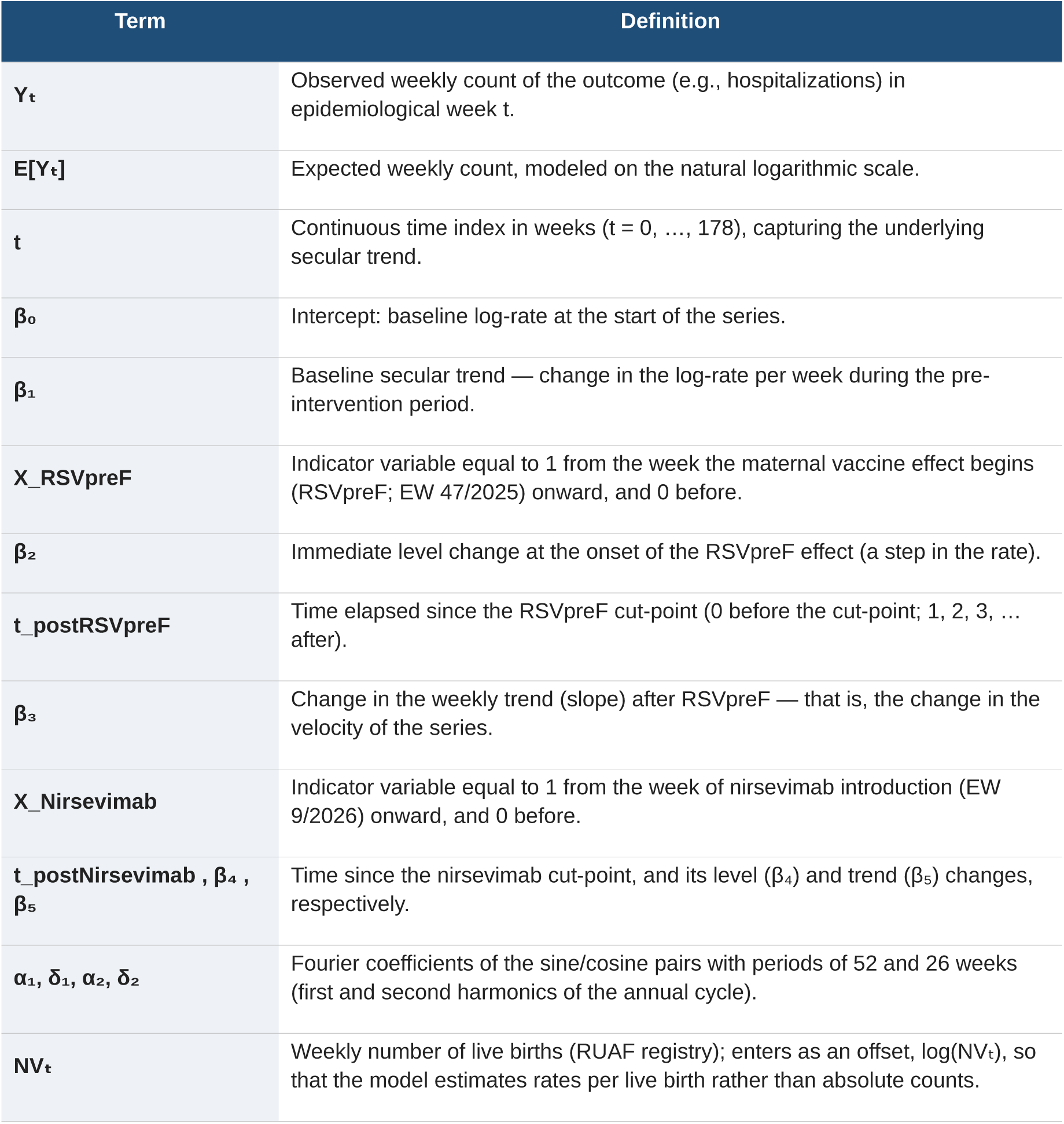
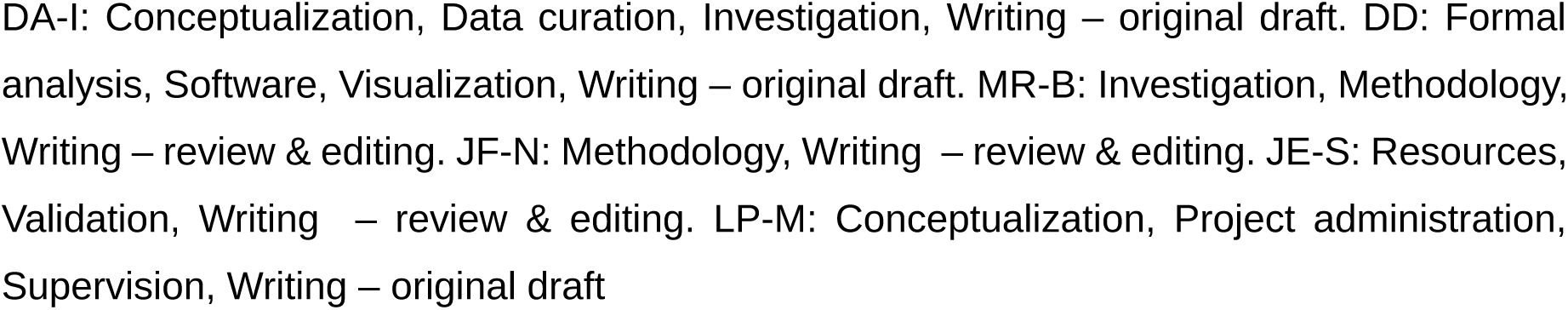
Definition of the terms in Equation 1.

| Term | Definition |
| --- | --- |
| $Y_t$ | Observed weekly count of the outcome (e.g., hospitalizations) in epidemiological week $t$. |
| $E[Y_t]$ | Expected weekly count, modeled on the natural logarithmic scale. |
| $t$ | Continuous time index in weeks ($t = 0, \dots, 178$), capturing the underlying secular trend. |
| $\beta_0$ | Intercept: baseline log-rate at the start of the series. |
| $\beta_1$ | Baseline secular trend — change in the log-rate per week during the pre-intervention period. |
| $X_{\text{RSVpreF}}$ | Indicator variable equal to 1 from the week the maternal vaccine effect begins (RSVpreF; EW 47/2025) onward, and 0 before. |
| $\beta_2$ | Immediate level change at the onset of the RSVpreF effect (a step in the rate). |
| $t_{\text{postRSVpreF}}$ | Time elapsed since the RSVpreF cut-point (0 before the cut-point; 1, 2, 3, ... after). |
| $\beta_3$ | Change in the weekly trend (slope) after RSVpreF — that is, the change in the velocity of the series. |
| $X_{\text{Nirsevimab}}$ | Indicator variable equal to 1 from the week of nirsevimab introduction (EW 9/2026) onward, and 0 before. |
| $t_{\text{postNirsevimab}}, \beta_4, \beta_5$ | Time since the nirsevimab cut-point, and its level ($\beta_4$) and trend ($\beta_5$) changes, respectively. |
| $\alpha_1, \delta_1, \alpha_2, \delta_2$ | Fourier coefficients of the sine/cosine pairs with periods of 52 and 26 weeks (first and second harmonics of the annual cycle). |
| $NV_t$ | Weekly number of live births (RUAF registry); enters as an offset, $\log(NV_t)$, so that the model estimates rates per live birth rather than absolute counts. |

DA-I: Conceptualization, Data curation, Investigation, Writing – original draft. DD: Formal analysis, Software, Visualization, Writing – original draft. MR-B: Investigation, Methodology, Writing – review & editing. JF-N: Methodology, Writing – review & editing. JE-S: Resources, Validation, Writing – review & editing. LP-M: Conceptualization, Project administration, Supervision, Writing – original draft

## Notes

### Competing Interest Statement

The authors have declared no competing interest.

### Author Declarations

The present study was approved by the Research Ethics Committee of the Secretaria Distrital de Salud de Bogota. The study consisted of a secondary analysis of information from routine public health surveillance systems and administrative records. No intervention, direct or indirect, was performed on patients, nor was there any contact with them. The information analyzed came from previously anonymized databases and was processed solely in aggregate form through weekly counts. At no time were individual data accessed or used that would allow the identification of persons; therefore, there was no risk to the confidentiality, privacy, or rights of the participants.

